# The Neurological Symptoms of ‘Long’ COVID-19: A Comparison with other Neurological Conditions and Implications for Healthcare Services

**DOI:** 10.1101/2020.07.21.20158816

**Authors:** T Wildwing, N Holt

**Author notes:** Joint authors: Tamar Wildwing, PhD candidate, Nicole Holt, PhD candidate. Correspondence to: Tamar Wildwing.

## Abstract

**Objective:** In response to the rapid spread of COVID-19, this paper provides health professionals with better accessibility to available evidence, summarising findings from a systematic overview of systematic reviews of the neurological symptoms seen in patients with COVID-19. Implications of so-called ‘Long Covid’ on neurological services and primary care and similarities with other neurological disorders are discussed.

**Methods:** Firstly, a systematic overview of current reviews of neurological symptoms of COVID-19 was conducted. Secondly the implications of these findings are discussed in relation to the potential effect on neurological services and the similarities in the experience of patients with COVID-19 and those with other neurological disorders.

**Results:** Twenty-nine systematic reviews were identified within seven databases, published between 11^th^ April 2020 and 27^th^ August 2020. The results indicated (so far), that COVID-19 exhibits two types of neurological symptoms; life threatening symptoms such as Guillain Barre Syndrome and encephalitis, and less devastating symptoms such as fatigue and myalgia. These so-called lesser symptoms appear to be emerging as longer-term for some sufferers and have been recently labelled Long Covid. When compared, these Long Covid symptoms are very similar to other neurological conditions such as Chronic Fatigue Syndrome (CFS) and Functional Neurological Disorder (FND).

**Conclusions:** Implications for neurological healthcare services in the UK may include longer waiting times and a need for more resources (including more qualified health professionals). There is also a possible change-effect on health professionals’ perceptions of other neurological conditions such as CFS and FND. Future research is recommended to explore changes in health professionals’ perceptions of neurological symptoms because of COVID-19.

## Introduction

As of 15^th^ September 2020, almost thirty million people worldwide have been affected by the novel coronavirus COVID-19.^1^ It has been found to cause neurological manifestations in up to 50% of patients.^2^ Several systematic reviews have been conducted into the neurological symptoms of COVID-19. However, some reviews focused purely on specific types of symptoms such as olfactory symptoms, whilst others focused on studies only in specific countries, for example at least a quarter of the studies within the reviews were undertaken in China. It was therefore deemed necessary firstly, to conduct a systematic overview of previously conducted systematic reviews to summarise the neurological symptoms of COVID-19 seen so far.

Secondly, it is becoming clear that COVID-19 affects many patients neurologically, and in some cases, symptoms are persisting. A recent BMJ webinar discussed the emergence of so-called ‘Long Covid’.^3 4^ This paper further discusses the concerns about the short and long-term effects of COVID-19 on medical and holistic neurological practice and where future resources will be required.

Thirdly, as COVID-19 is a new disease, the potential similarities with other neurological conditions have not yet been explored in the literature. The authors of this paper identified that many of the neurological symptoms of COVID-19 appear similar to the symptoms of FND. In addition, the BMJ webinar and recent articles^4^ 5 discussed the similarities between so-called ‘Long Covid’ and CFS, as it has become increasingly noticeable that symptoms are similar. Whilst FND is generally regarded as a psychological disorder, the authors of this paper argue that the emerging discussions about supporting patients with ‘Long Covid’^6 7^ should relate to other neurological disorders such as FND and CFS, questioning how these conditions are judged and how treatments are resourced for them as well as for ‘Long Covid’.

Neurological services in the UK offer diagnosis and treatment to patients with disorders of the nervous system.^8^ This research is therefore aimed at health professionals and commissioners in the field of neurology and in primary care in the UK. It aims to inform future service provision for those who develop long-term neurological symptoms due to COVID-19; as well as exploring the effect of COVID-19 on health professionals’ perceptions towards those with symptoms of FND.

## Methods

A systematic overview of current systematic reviews was conducted to explore the potential impact of the longer-term neurological symptoms of COVID-19. The review protocol was not previously registered. The implications of the findings of this overview were discussed, including potential effects on neurological and primary care services and on perceptions towards patients with other neurological disorders such as CFS and FND.

To conduct the systematic overview of systematic reviews, the recommendations outlined in the Preferred Reporting Items for Systematic Reviews and Meta-Analyses (PRISMA) statement were followed (Supplementary Material 1).^9^ Following these recommendations reduces the risk of bias or selective reporting and demonstrates transparency in the process conducted. The following databases were searched between December 2019 and June 2020: ‘PubMed Central’, ‘Cochrane Database of Systematic Reviews,’ ‘Ovid’ ‘ScienceDirect’ ‘Biomed Central’ ‘BMJ’ and SAGE Journals. The following keywords within the title or abstract were used to conduct each search: ‘coronavirus’, ‘COVID-19’, ‘SARS-COV-2’, ‘neurological’, ‘nervous’ and ‘review’. For example, PubMed Central was searched using the following search terms: (((COVID-19 OR SARS-COV-2 OR coronavirus[Title]) OR (COVID-19 OR SARS-COV-2 OR coronavirus[Abstract])) AND ((neuro* OR nervous[Title]) OR (neuro* OR nervous[Abstract])) AND ((review[Title] OR (review[Abstract]))). Limitations: Date: 01/12/2019-01/09/2020. Only articles published in academic journals in English were retrieved. Reference lists of retrieved articles were also searched to ensure literature saturation.

The inclusion criteria consisted of systematic reviews only relating to neurological symptoms seen in patients with COVID-19 since 1^st^ December 2019. Both authors participated through each step of the review independently (screening, eligibility and inclusion). Reviews were screened for relevancy against the inclusion criteria within title and abstract. Full-text reports for all potentially relevant reviews were obtained, including those where there was any uncertainty. The authors screened the full-text reports for relevancy and resolved any disagreement through discussions. Neither of the authors were blind to the journal titles, the authors’ of the reviews or institutions. The Critical Appraisal Skills Programme (CASP) checklist for systematic reviews (2018) was used to establish the quality of each review included within this systematic overview.^10^ All included reviews were deemed high quality; 23 reviews met nine of the ten CASP criteria, five reviews met eight of the criteria and one review met seven of the criteria. All reviews were read in full by both authors. Microsoft Excel was used to compile a list of all the included reviews and the neurological symptoms mentioned in each. This list was completed and checked by both authors.

Next, neurological symptoms of COVID-19, CFS and FND were placed into a table to compare the symptoms, which led to a consideration of the implications for neurological healthcare services and primary care in the UK, and to the possible change-effect on perceptions towards CFS and FND.

## Results

### A systematic overview of systematic reviews of neurological symptoms of COVID-19

PRISMA guidelines were followed for the search strategy of the systematic overview as seen in Figure 1. The database search identified 438 papers, with a further 13 papers identified within the reference lists of the included reviews. From a total of 451 papers, 381 were excluded after title and abstract review for relevance. The remaining 56 papers were reviewed in full. 27 of these were excluded as they did not meet the inclusion criteria (date, study design or methodology). 29 systematic reviews met the inclusion criteria and were included in this overview 11 12 13 14 15 16 17 18 19 20 21 22 23 24 25 26 27 28 29 30 31 32 33 34 35 36 37 38 39

**Figure 1:**
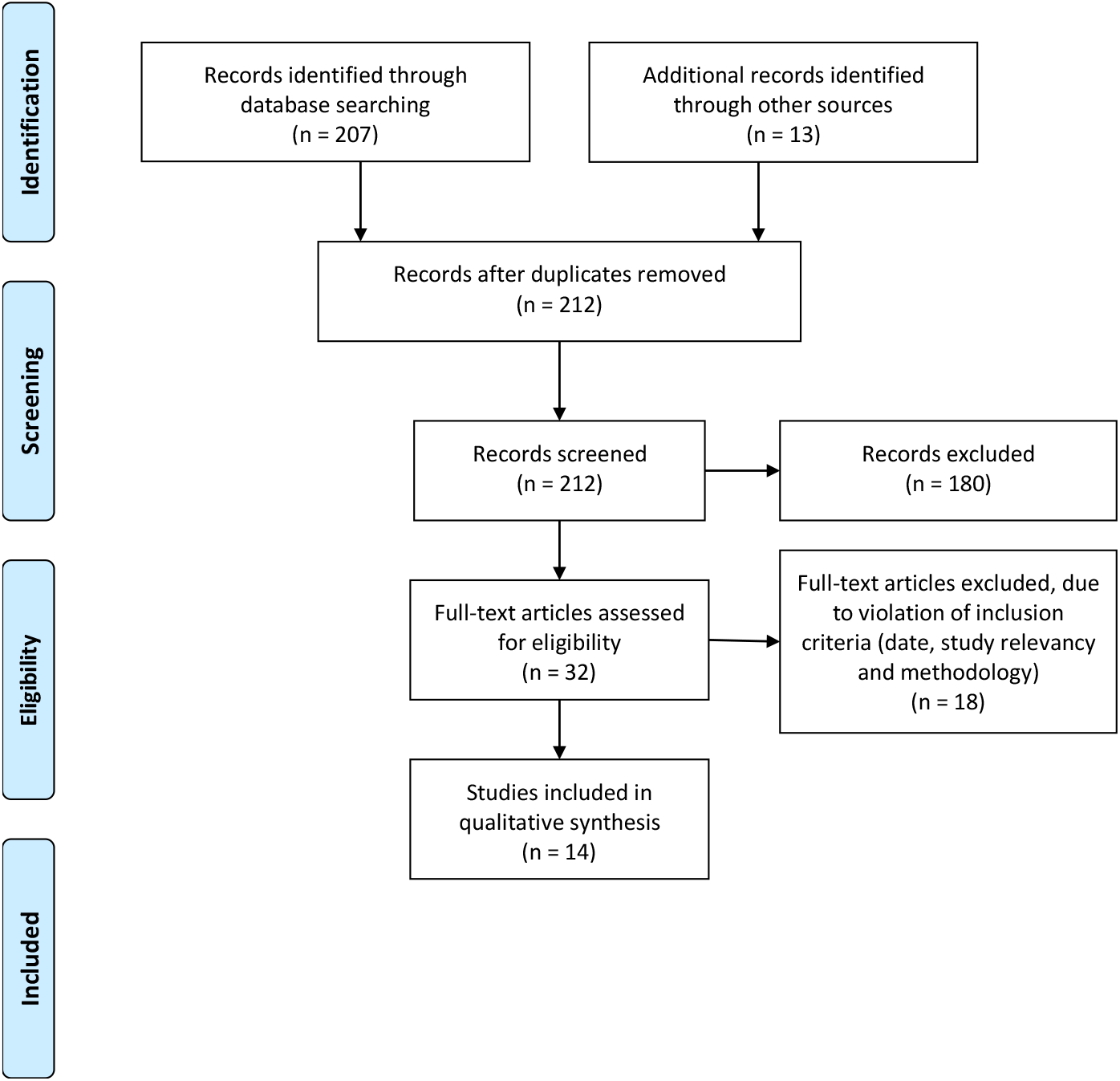
Preferred reporting items for systematic reviews and meta-analyses (PRISMA) flow diagram.

The 29 systematic reviews were published between 11^th^ April 2020 and 27^th^ August 2020. The characteristics of the reviews can be seen in Table 1; the number of relevant studies reviewed within each review, the number of participants in each relevant study and the aims of each study. As mentioned, the CASP checklist was utilised to check for risk of bias in each review.

**Table 1:** Characteristics of the reviews

| **First author** | **No. of studies referenced\*** | **No. of participants\*** | **Aim of review** |
| --- | --- | --- | --- |
| Abdullahi<sup>11</sup> | 60 | 10314 (11069) | [T]o summarize the evidence on the neurological and musculoskeletal symptoms of the disease. This may help with early diagnosis, prevention of disease spread, and treatment planning. |
| Abu-rumeileh<sup>12</sup> | 52 | 58 (73) | [T]o provide a comprehensive and updated overview of all case reports and series of COVID-19-related GBS to identify predominant clinical, laboratory, and neurophysiological patterns and to discuss the possible underlying pathophysiology. |
| Agyeman<sup>13</sup> | 24 | 8438 | To estimate the prevalence of olfactory and gustatory dysfunctions (OGDs) among patients infected with novel coronavirus disease 2019 (COVID-19).' |
| Almqvist<sup>14</sup> | 41 (378) | 11509 (14418) | [T]o systematically summarize neurological and neuroimaging manifestations of all known HCoVs in order to provide possibilities to predict short- and long-term neurological complications of COVID-19. |
| Asadi-Pooya<sup>15</sup> | 6 (8) | 765 (not stated) | [To] 'discuss the evidence on the occurrence of central nervous system (CNS) involvement and neurological manifestations in patients with COVID-19.' |
| Ceravolo<sup>16</sup> | 18 (36) | 346 (not stated) | [T]o gather and present the current evidence informing rehabilitation of patients with COVID-19 and/or describing the consequences due to the disease and its treatment. |
| Collantes<sup>17</sup> | 47 (49) | 6094 (6335) | [T]o determine the neurological manifestations and complications, including laboratory findings, and outcomes among patients with COVID-19 101 infection. |
| Correia<sup>18</sup> | 4 (7) | 329 (409) | To describe the main neurological manifestations related to coronavirus infection in humans.' |
| Di Carlo<sup>19</sup> | 19 | 6958 (12157) | The primary objective of this systematic review was to analyse the overall rate of neurologic symptoms among COVID-19 patients. Secondary objective was to compare the results between patients with severe and non-severe infection. |
| Dinakaran<sup>20</sup> | 31 (12) | 2552 (399) | [To] 'report the available evidence of neuropsychiatric morbidity during the current COVID-19 crisis. The authors also discuss the postulated neuronal mechanisms of the corona virus infection sequelae.' |
| Ghannam<sup>21</sup> | 43 (42) | 314 (82) | To clarify the neurological complications of SARS-CoV-2 infection including the potential mechanisms and therapeutic options. |
| Katal<sup>22</sup> | 21 (28) | 681 (not stated) | [S]ystematically reviewed the available imaging findings of patients diagnosed with neurological symptoms associated with coronavirus infections. |
| Leonardi<sup>23</sup> | 9 (29) | 510 (not stated) | 29 papers related to neurological manifestations associated with COVID-19 were examined. |
| Montalvan<sup>24</sup> | 20 (67) | 749 | To review the neurological aspects of SARS-cov2 and other coronavirus, including transmission pathways, mechanisms of invasion into the nervous system, and mechanisms of neurological disease.' |
| Nepal<sup>25</sup> | 37 | 3152 | [T]o inform and improve decision-making among the physicians treating COVID-19 by presenting a systematic analysis of the neurological manifestations experienced within these patients. |
| Pinzon<sup>26</sup> | 33 | 7113 (7559) | To conduct a systematic review and meta-analysis on the neurologic characteristics in patients with COVID-19. |
| Rogers<sup>27</sup> | 12 (72) | 1048 (3559) | [T]o assess the psychiatric and neuropsychiatric presentations of SARS, MERS, and COVID-19.' |
| Scoppettuolo<sup>28</sup> | 40 (42) | 1640 (903) | [T]o provide a clinical approach of SARS-CoV-2 neurological complications based on the direct or indirect (systemic/immune-mediated) role of the SARS-CoV-2 in their genesis.' |
| Sharifan<sup>29</sup> | 177 (208) | 17595 (not stated) | To summarize available information regarding the potential effects of different types of CoV on the nervous system and describes the range of clinical neurological complications that have been reported thus far in COVID-19. |
| Taherifard<sup>30</sup> | 21 (22) | 489 (57) | [T]o systematically review the neurological complications in patients with SARS-CoV-2 infection and the methods used to diagnose both neurological complications and coronavirus infection. |
| Tan<sup>31</sup> | 37 (39) | 4720 (135) | [T]o characterize the clinical characteristics, neuroimaging findings, and outcomes of AIS [Acute ischemic stroke] in COVID-19 patients |
| Tsai<sup>32</sup> | 36 (142) | 3116 (not stated) | Review and integrate the neurologic manifestations of the Coronavirus Disease 2019 (COVID-19) pandemic, to aid medical practitioners who are combating the newly derived infectious disease.' |
| Tsivgoulis<sup>33</sup> | 13 (not stated) | 1641 (not stated) | [T]o present the neurological manifestations associated with SARS-CoV-2 infection and COVID-19. ... We also evaluated the impact of the COVID-19 pandemic on the health care of neurological patients.' |
| Uncini<sup>34</sup> | 33 | 21 (42) | [T]o clarify the clinical and electrophysiological phenotype, to discuss, on the basis of the available data, whether the disease mechanism could be parainfective or postinfective and to speculate on the possible pathogenesis. |
| Vonck<sup>35</sup> | 21 (20) | 3575 (3423) | [T]o perform a review to describe neurological manifestations in patients with COVID-19 and possible neuro-invasive mechanisms of Sars-CoV-2.' |
| Wang<sup>36</sup> | 41 | 4345 (not stated) | [T]o systematically collect and investigate the clinical manifestations and evidence of neurological involvement in COVID-19. |
| Werner<sup>37</sup> | 14 (not stated) | 3351 (not stated) | [To] conduct ... a review of the reported data for studies concerning COVID-19 pathophysiology, neurological manifestations, and neuroscience provider recommendations and guidelines.' |
| Whittaker<sup>38</sup> | 32 (31) | 2582 (2504) | [R]eports are emerging of the virus' effects systemically, including that of the nervous system. A review of all current published literature was conducted' |
| Wilson<sup>39</sup> | 10 | 330 (not stated) | [T]o evaluate and summarize the current status of the COVID-19 literature at it applies to neurology and neurosurgery. Neurological symptomatology, neurological risk factors for poor prognosis, pathophysiology for neuroinvasion, and actions taken by neurological or neurosurgical services to manage the current COVID-19 crisis are reviewed.' |

\*Many reviews referenced the same papers, in some cases reviews stated a different number of studies than were referenced within the review, some reviews did not state no. of participants, (no. in brackets is total no. of studies/participants where the review included differing no. of studies from this review), when reviews differed in their report of participants nos. within the same study a consensus was assumed

As can be seen in Table 1, some of the reviews explored issues outside the scope of this review. Within scope of this review, 417studies relating to neurological symptoms of COVID-19 were reviewed within the 29 reviews. Most of the reviews included the same studies. For instance, Mao et al. (2020) was included in 21 reviews.^40^ Whilst numbers of participants were not always stated, at least 43,166 participants were included across the 417 studies reviewed.

Table 2 shows the country where each reviewed study took place. As can be seen, a quarter of the studies took place in China (107 studies with a total of 17,855 participants). Italy, South Korea, Spain, the UK and the USA researched more than 2000 participants in each country. This gives an overview of the countries in which neurological symptoms of COVID-19 are under investigation, although it was not determined whether all participants were within the country stated. There may also be publication bias as this review excluded reviews that were not in English.

**Table 2:** Countries where studies took place

| **Country** | **No. studies in total** | **No. participants in total (where stated)** | **No. studies where no. of participants were not stated (within reviews)** |
| --- | --- | --- | --- |
| Austria | 1 | 1 |  |
| Belgium | 2 | 47 |  |
| Brazil | 1 | 1 |  |
| Canada | 1 | u/k | 1 |
| China | 107 | 17855 | 10 |
| Europe | 2 | 1837 |  |
| France | 17 | 985 | 2 |
| France/Spain | 1 | 1 |  |
| France/Switzerland | 1 | 6 |  |
| Germany | 8 | 185 |  |
| Hong Kong | 1 | 50 |  |
| India | 3 | 66 |  |
| Iran | 13 | 75 | 4 |
| Israel | 1 | 42 |  |
| Italy | 31 | 2260 | 5 |
| Japan | 2 | 2 |  |
| Morocco | 1 | 1 |  |
| Netherlands | 4 | 447 | 1 |
| Poland/USA | 1 | u/k | 1 |
| Romania | 1 | 126 |  |
| Saudi Arabia | 2 | 2 |  |
| South Korea | 4 | 3275 |  |
| Spain | 16 | 5377 | 6 |
| Sudan | 1 | 1 |  |
| Sweden | 1 | 1 |  |
| Switzerland | 4 | 12 | 1 |
| Thailand | 1 | u/k | 1 |
| Turkey | 7 | 90 |  |
| United Arab Emirates | 2 | 1 | 1 |
| United Kingdom | 12 | 2369 | 2 |
| USA | 39 | 7724 | 10 |
| Country not stated | 129 | 327 | 67 |
| **Total: 31 different countries** | **417 novel studies** | **at least 43166 participants** | **112 studies where participants could not be counted** |

Throughout the 417 studies, 22 neurological symptoms were described. These are summarised in Table 3. As can be seen, neurological manifestations of COVID-19 include catastrophic symptoms such as cardio-vascular disease (CVD), encephalitis and Guillain-Barre Syndrome, which are understandably hugely concerning and have therefore generated intense discussion and research. COVID-19 has also been found to cause symptoms such as fatigue, dizziness, ataxia, dysphagia and headache, which though more benign, can be disabling if they become chronic.

**Table 3:**
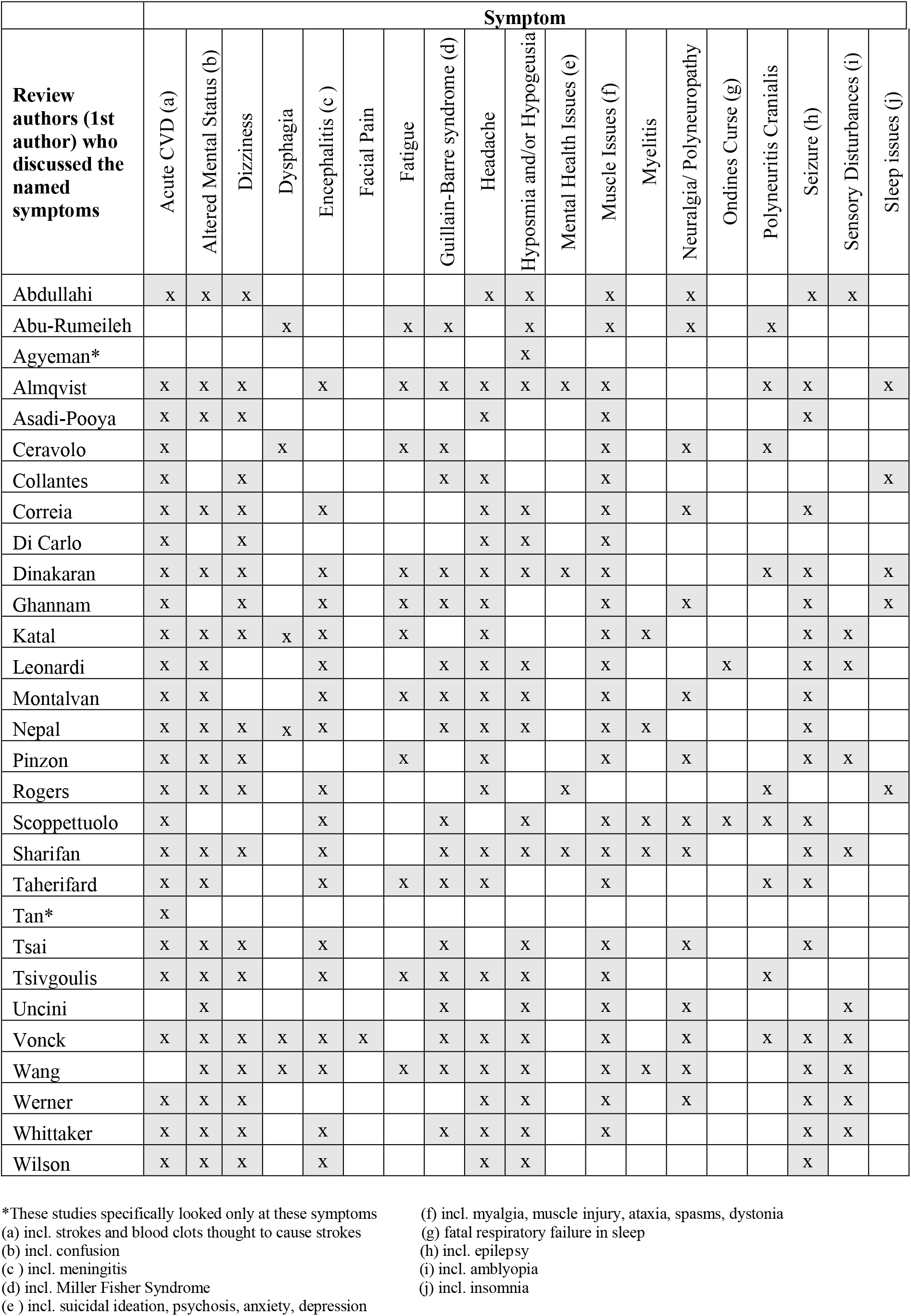
Neurological symptoms of COVID-19

### A comparison between the neurological symptoms of COVID-19 and the most common symptoms of FND and CFS

The most common symptoms experienced by people with FND and those with CFS are compared with the neurological symptoms of COVID-19 in Table 4. The symptoms of FND and of CFS were collated from key NHS sources and DSM-5. ^41 42 43 44^ Neurological symptoms of COVID-19 include catastrophic symptoms requiring emergency care such as stroke, brain haemorrhage, encephalitis and Guillain-Barré Syndrome. However, 15 other neurological symptoms were shown to be similar in COVID-19 and FND and/or CFS (shaded in grey within Table 4). In fact, every non-catastrophic symptom of COVID-19, including the much-researched symptom of hyposmia, is also described as a symptom of FND and/or CFS. Furthermore, most of the symptoms of FND and CFS, have been experienced by some people with COVID-19 neurological symptoms, particularly those with Long Covid.

**Table 4:**
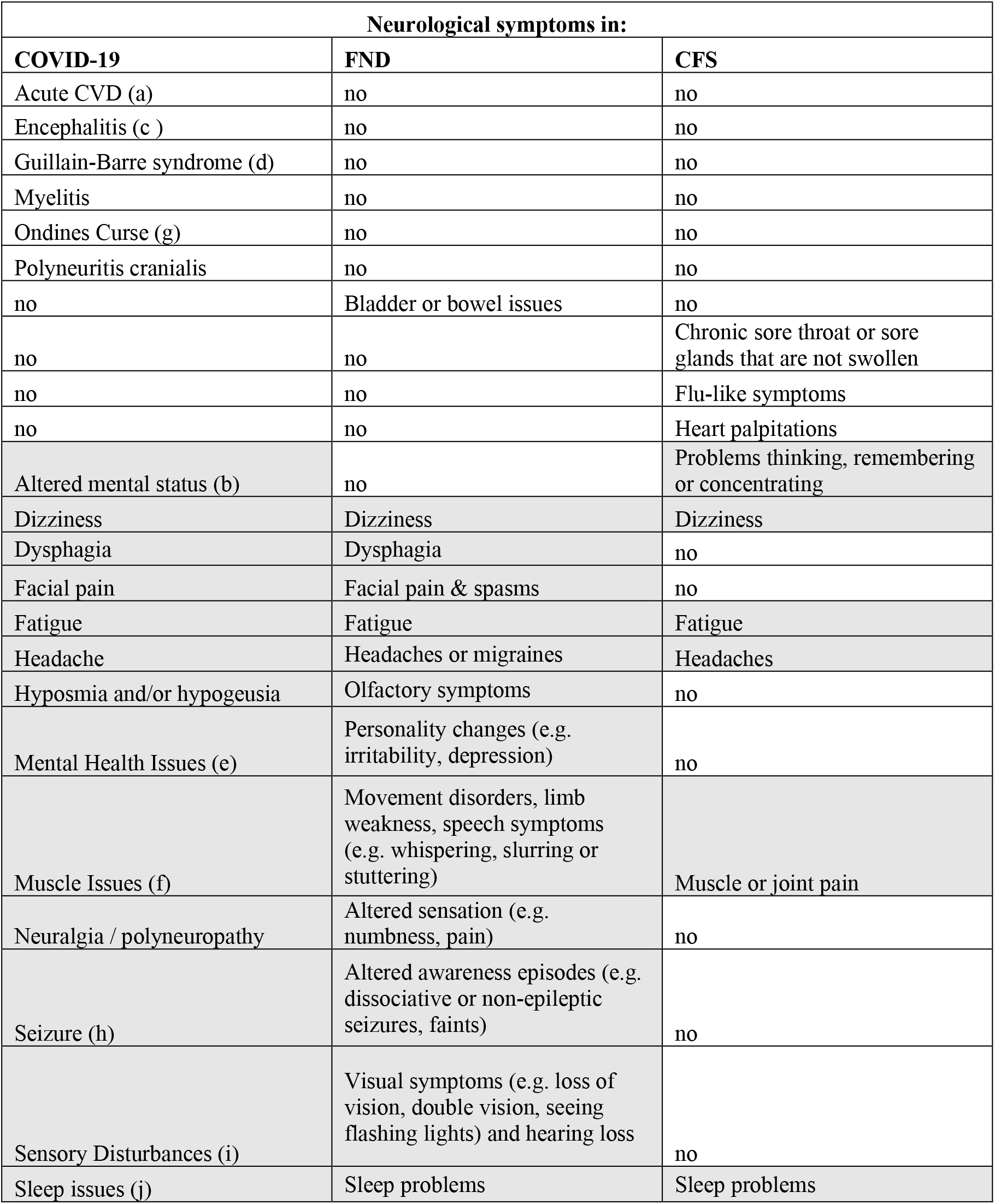
Comparison between neurological symptoms in COVID-19, FND and CFS

## Discussion

From these results, this paper argues three main points: 1) COVID-19 appears to be becoming chronic for some patients, presenting similar neurological symptoms to FND and to CFS; 2) the long-term neurological effects of COVID-19 may impact neurological and wider health care services; 3) the neurological symptoms seen in COVID-19 may (and should) affect health professionals’ perceptions of FND and CFS.

### 1) Long Covid, FND and CFS present similar neurological symptoms

Long Covid, FND and CFS present similar neurological symptoms; every non-catastrophic symptom of COVID-19 (except for hypogeusia) is described as a symptom of FND and/or CFS. In addition, every symptom of FND (except bladder and bowel issues), and of CFS, (except flu symptoms and heart palpitations) have been experienced by some people with COVID-19. Notably, the main symptoms of Long Covid are increasingly similar to symptoms of CFS (fatigue, inability to concentrate, myalgia, headache). ^3^

### 2) The impact of these findings on neurological healthcare services and primary care

Echoing concerns raised recently by the British Medical Association,^7^ the findings of this review highlight the impact that short and long-term neurological symptoms of COVID-19 may have on current health services. Symptoms seen in Long Covid such as facial pain, muscle issues, neuralgia, fatigue and insomnia, may become long term and disabling, requiring sustained support from healthcare services such as pain, fatigue and sleep clinics, neurological services and primary care. This is supported by other research which suggests rehabilitation required by patients following COVID-19 infection may be ‘very much along the same lines as existing services but with double the demand’.^45^ Consultant-led neurological services such as chronic fatigue clinics and headache clinics are already overstretched with a shortage of neurology consultants and long waiting lists.^46^ Neurological symptoms of Long Covid may increase demand for these clinics and may indicate a need for more qualified health professionals and specialists in neurology. The effects of COVID-19 on these services is hard to predict, as the neuropathy, myopathy and sensory deficits of SARS resolved within three months of recovery.^32^ However, as COVID-19 appears to be becoming Long Covid for up to 10% of patients,^3^ support is likely to be required, potentially for a significant number of people, if their symptoms do not resolve spontaneously.

Additionally, COVID-19 is causing a wider impact on patient populations. For example, COVID-19 has affected delivery of health care services, through reduction in use of emergency services during the peak of the pandemic in the UK as patients were worried about contracting the virus from hospitals.^47^ There is evidence that some patients with long term conditions have improved their self-care techniques, such as better use of medication and alternative therapies such as physiotherapy, cognitive behavioural therapy, and exercise. On the other hand, most outpatient appointments and elective surgeries were postponed, leading in some cases to deaths as an indirect result of COVID-19. ^48^ There are now questions about whether there will be a rebound in demand, potentially overwhelming NHS services, or whether the reduction in demand can be sustained. Combined with the previously mentioned potential increase in demand for neurological services, there is likely to be wide reaching financial implications. This research is therefore useful for aiding future patient management while helping to develop policies for response to COVID-19 and its critical outcomes.

### 3) The effect of COVID-19 on perceptions of FND and CFS

Key sources differ in their description of FND symptoms, indicating that there is no comprehensive list of all FND symptoms. ^41 42 43 44^ It is likely that this contributes to health professionals’ uncertainty in diagnosing FND.^49^ In addition, patients with both FND and CFS have experienced many years of scepticism from health professionals^5^ with negative consequences, including lack of support and poor access to services often contributing to poor mental health. ^50^ In addition, lower value is given to health professionals’ role in managing poorly defined symptoms.^51^ FND and CFS, alongside other ambiguous uncertain conditions (e.g. fibromyalgia), are considered amongst the lowest conditions on the hierarchy of importance of conditions.^52^

Conversely, COVID-19 can be quickly and easily diagnosed with a test (leaving aside the possibilities of false results). The authors of this research are concerned that within neurological services, priority may be given to patients who have had COVID-19, who may actually experience some level of prestige because they have survived a disease feared by all, and any neurological symptoms they experience might be automatically accepted, extensively researched and supported, at detriment to other neurological conditions. There is already evidence of this skewing of services, as large sums of money and research at pace are contributing to a greater understanding of long-term symptoms of COVID-19,^53^ and the NHS has declared its intention to provide a COVID-19 rehabilitation service.^13^ Also, there is indication that the presence of Long Covid is unquestionably ‘believed’ by health professionals, as doctors and other professionals have written about their experiences with it^5^ and are recommending each other’s papers.^3^

However, some patients with emerging Long Covid have similar experiences to those with less accepted conditions, particularly those who were unable to get a test early in the pandemic^54^. Patients feel doctors dismiss their symptoms, they feel desperate, and disagree with diagnoses of anxiety. As Garner (2020) explained, “Doctors need to stop diagnosing this as anxiety. We have messed up before, lets’ not do it again with long term covid-19 illness.”^5^

It is important to highlight the similarities between symptoms of Long Covid and other conditions, providing education for health professionals, informing future practice and illustrating the need for more funding for neurological services to meet increased demand. Whilst research and services for COVID-19 are gaining funding, there is a lack of funding and research into understanding, treatment and support for those who suffer from FND and CFS.^55^ FND and CFS are likely to have lower prestige than COVID-19, however the realisation that COVID-19 causes neurological symptoms similar to FND and CFS may lead to a potential shift in perceptions towards these conditions. They may be taken more seriously, and more funding may be made available for appropriate neurological services unrelated to the cause of the symptoms.

## Limitations

Scientific reports centred on neurological effects of COVID-19 are still scarce, and risk of publication bias is high. For example, within the reviews included in this research, a quarter of the studies were undertaken in China (107) and a further quarter in Europe (99). Only two studies were conducted in the African continent and none were conducted in the South American continent. Despite the status of COVID-19 as a pandemic, research into the neurological effects of COVID-19 so far has not been conducted worldwide.

This systematic review is grounding its results on previous reviews’ findings; thus, it is difficult to assess how reliable some of these results can be, for example many of the reviews (67) did not state the source of their findings. Quality assessment has however been undertaken as described earlier and the reviews deemed high quality. Retrospective and prospective studies of larger cohorts are necessary to correctly assess nervous system involvement, which has not been possible yet for COVID-19 as it is a very new disease and it is unclear how much it mimics other coronaviruses.

## Conclusion and implications for future research

There is an array of evidence to show that COVID-19 causes neurological symptoms, and although it is difficult to ascertain how long-term the symptoms may become, there is increasing evidence of the presence of Long Covid, symptoms persisting beyond three months. Although this paper is primarily UK focused, these concerns are likely to be similar in other countries. This research collates the evidence so far and provides insight into the neurological effects of COVID-19 in relation to FND symptoms. Concerns about the potential impact of these findings on the delivery of neurological and wider healthcare services are considered alongside the potential effect COVID-19 may have on perceptions of neurological symptoms, particularly those relating to FND and CFS. Further research is recommended to explore whether the neurological symptoms of COVID-19 will improve acceptance and understanding of FND or whether this will worsen the experience for those who suffer from FND. Further thought for future planning of health care resources also needs to be taken into consideration, in light of this pandemic.

## Data Availability

All references referred to in the manuscript are published journal articles or accessible webpages.

